# Frailty trajectories in the oldest old: Is the level or the rate of change more predictive of mortality?

**DOI:** 10.1101/2021.10.01.21264405

**Authors:** Erwin Stolz, Hannes Mayerl, Emiel O. Hoogendijk

## Abstract

**BACKGROUND:** It is unclear whether frailty index (FI) change captures mortality risk better than and independently of the current FI level, i.e. whether a regular FI assessment among older adults provides additional insights for mortality risk stratification or not.

**METHODS:** We used data from the LASA 75-PLUS-study, which monitored health among 508 older adults (75+) between 2016-2019 every 9 months. Joint models for longitudinal and time-to-event data were used to assess the impact of both current FI and within-person FI change during the last year on mortality risk.

**RESULTS:** 20% of the participants died during 4.5 years of follow-up. Adding within-person FI change to the current FI model improved model fit and it showed that FI increases during the last year were associated with a strong increase in mortality risk. Consequently, the effect of the current FI decreased considerably and became statistically non-significant.

**CONCLUSIONS:** The rate of FI change was more important than the current FI level for short-term mortality prediction among the oldest old, which highlights the benefits of regular frailty assessments.

## 1. Introduction

Frailty describes a state of vulnerability to stressors among older adults due to a cumulative decline in multiple physiological systems[1] which has implications for both clinical practice and public health[2]. One of the two main conceptualizations[3] of frailty is the frailty index (FI) based on the health deficit accumulation approach[4,5], which has been shown to consistently predict mortality among older adults[6]. Most of these studies, however, rely on static, one-time assessments, although it has been shown that individuals’ FI change over time[7–10]. A few studies[11–17] have assessed the relationship between frailty changes or trajectories and mortality, generally finding FI increases or steeper trajectories to be associated with a higher mortality risk. However, mixed results have been found with regard to the question whether FI change captures mortality risk better than and independently of the current (most recent) FI observation[13,17]. In other words, it is unclear if the current FI level is sufficient or whether a regular FI assessment provides additional insights for mortality risk stratification. In the current study, we use more intensive data (5 assessments with 9-month intervals) and a more advanced statistical method (joint models for longitudinal and time-to-event data) than previous studies to answer this question.

## 2. Methods

### Data

Data came from the Longitudinal Aging Study Amsterdam (LASA), an ongoing Dutch study of older adults based on a nationally representative sample[18]. Specifically, we used data from the ancillary study of the oldest old (75-PLUS-study) in LASA, which monitored health and functional changes among 510 older adults (75+) in up to five consecutive face-to-face interviews every 9 months between 2015 and 2019[19].

### Variables

#### Mortality

Based on register data from municipalities, mortality was followed-up until March 2020 (100% complete). A time-line was created beginning with the first interview until either the time of the event or the end of follow-up (up to 4.5 years).

#### Frailty index

Frailty was operationalized with a FI following a standard procedure[20,21]. Specifically, the FI was calculated from 32 health deficits in each wave (Appendix 1). The values of the health deficits were summed-up and divided by the number of valid health deficits (e.g. 6/32 = 0.19). In total, this amounted to a final sample size of 508 participants (two participants had >20% missing data) who provided 2059 repeated observations over 3.5 years.

### Statistical analysis

To predict mortality, we used joint models for longitudinal and time-to-event data[22,23], an advanced statistical approach where the effect of a time-varying predictor (frailty) on the time-to-event outcome (mortality) is estimated based on its longitudinal characteristics captured with mixed regression models. The best model fit for the mixed model was obtained with a non-linear approach using cubic splines at both the population- and individual-level. The survival regression submodel was formulated as:

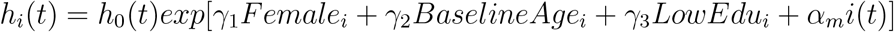

where the risk of mortality *h* for individual *i* at time *t* depends on the baseline hazard function (*h*_0_) estimated with a cubic spline, the regression coefficients (*γ*_1*-*3_) of the time-constant sociodemographics, and the association parameter(s) (*α*). In the first model (JM1), *α*_1_ refers to the current value parametrization, where the ‘true’ FI value based on the individualized growth curves at *t* is used as a time-varying predictor. In the second model (JM2), we added *α*_2_ which quantifies the association between the within-person FI trajectory during the last year before *t* and the risk of mortality. The Watanabe-Akaike Information Criterion (WAIC) and the Deviance Information Criterion (DIC) based on the marginal log-likelihood were used for model comparison. Both criteria[24] correct for the number of parameters, and smaller values indicate better model fit. Discrimination and calibration were evaluated with time-varying area-under-the-curve (AUC) values and Brier scores using FI observations up to year 2 (474 subjects still at risk) and 1- and 2-year prediction windows. Joint models were fitted using R-package JMbayes2 (v0.1-8). All calculations were done in R (v4.0.3).

## Results

At baseline, participants’ average age was 81.9 (SD=5.3, range=75-99) years, 60.4% of the sample were women, and 46.3% had only completed elementary or lower vocational education. The mean/median FI at baseline was 0.23/0.21 (SD=0.13/IQR=0.17, range=0-0.67). During the 4.5-year follow-up, 19.9% of the participants died after a mean of 2.5 years (SD=0.9, range=0.8-4.3). Figure 1 shows the mean FI trajectories by event group, indicating that participants who died had higher FI values on average and steeper FI increases than survivors. Results from the longitudinal submodel (JM2 in Table 1) indicate that non-linear FI growth was higher among those with higher initial FI levels and that earlier FI increases correlated with later FI increases.

**Table 1:** Results from joint longitudinal and time-event models

|  | JM1 | JM2 |
| --- | --- | --- |
| LONGITUDINAL OUTCOME | $\beta/b$ (CI-95) | $\beta/b$ (CI-95) |
| Population-level (fixed) effects |  |  |
| Intercept | 22.70 (21.31-24.07) | 23.03 (21.57-24.47) |
| Time (Spline 1/3) | -1.10 (-2.31-0.11) | 0.65 (-0.46-1.64) |
| Time (Spline 2/3) | 4.73 (3.29-6.18) | 3.64 (2.34-4.99) |
| Time (Spline 3/3) | 5.28 (4.10-6.45) | 5.55 (4.40-6.71) |
| Individual-level (random) effects |  |  |
| SD Intercept | 12.14 | 11.83 |
| SD Time (Spline 1/2) | 6.85 | 5.84 |
| SD Time (Spline 2/2) | 6.92 | 4.32 |
| Corr. Intercept*Time (Spline 1/2) | 0.03 | 0.31 |
| Corr. Intercept*Time (Spline 2/2) | 0.22 | 0.35 |
| Corr. Time (Spline 1/2)*Time (Spline 2/2) | 0.13 | 0.65 |
| EVENT PROCESS | HR (CI-95) | HR (CI-95) |
| Female ($\gamma_1$) | 0.69 (0.43-1.09) | 0.40 (0.16-1.00) |
| Age ($\gamma_2$) | 1.04 (1.01-1.07) | 1.11 (1.05-1.19) |
| Low education ($\gamma_3$) | 0.99 (0.75-1.30) | 1.08 (0.56-2.09) |
| Current FI ($\alpha_2$) | 1.03 (1.02-1.04) | 1.01 (0.97-1.05) |
| FI change (last year) ($\alpha_1$) | - | 6.61 (4.31-10.51) |
| MODEL FIT |  |  |
| WAIC | 15073 | 15023 |
| DIC | 15085 | 14963 |

Number of participants = 508, number of repeated observations of frailty index = 2,059, number of events = 101 (19.9%). Frailty index was multiplied with 100 so that hazard ratios refer to 0.01 FI differences. $\beta$ = population-level (fixed) effects from longitudinal submodel, $b$ = individual-level (random) effects from longitudinal submodel, HR = hazard ratio of survival submodel, $\gamma$ = effects of time-constant predictors, $\alpha$ = effects of time-varying predictors, CI-95 = 95% credible intervals, WAIC = Watanabe-Akaike Information Criterion, DIC = Deviance Information Criterion. Model fit with MCMC estimation procedure with 3 chains, 50000 iterations per chain, 10000 iterations burn-in, and thinning by 1. All $\hat{r}$ -values < 1.1.

**Figure 1:**
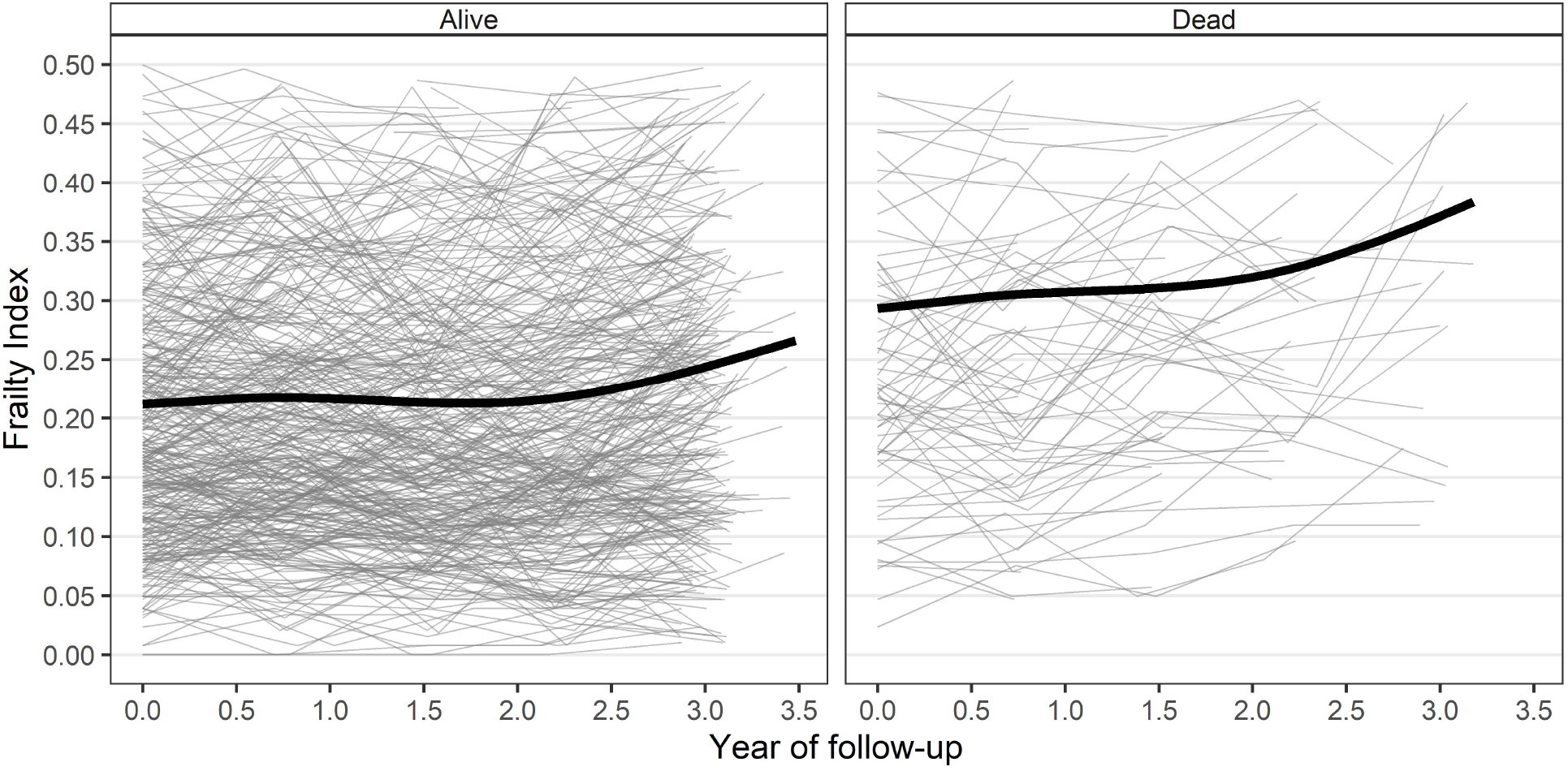
Frailty index trajectories by event status Figure shows smoothed average trajectory of frailty index by event group (thick black line) and individual frailty index measurements (thin grey lines).

The results from the survival submodel with only the current value parametrization (JM1) indicate that – adjusted for sociodemographics – a 0.01 higher current FI was associated with a statistically significant (p<0.001) increase of the mortality risk by 3%. Model comparison, however, indicates that the second joint model (JM2), which adds the time-dependent slope parametrization, provided better predictive performance (smaller WAIC and DIC). In the second model, a one unit steeper slope of the FI trajectory during the last year was associated with a strong increase in mortality risk. Adding the within-person FI change as a predictor of mortality decreased the effect size of the current FI value considerably (from 3% to 1%) and rendered it statistically non-significant (p-value=0.660). The second model provided adequate discrimination and calibration for mortality prediction: 1-year AUC/Brier=0.76/0.06, 2-year AUC/Brier=0.71/0.12.

## Discussion

In this study, we assessed whether the within-person FI rate of change predicts mortality above and beyond the current FI level. In other words, we wanted to know whether it also matters for risk prediction how fast an older person arrived at a specific current FI level. Based on an average of four repeated FI measurements every nine months among a population-representative sample of people aged 75+, we found the rate of FI change to be more important than the current FI level for short-term mortality prediction. Adjusting for last year’s rate of FI change greatly reduced the predictive effect of the current FI level for mortality. Our results suggest that FI levels changed considerably during the observation period, and that the FI should be assessed regularly among the oldest old to predict the risk of death. Such regular frailty assessments could be integrated in annual health check-ups or derived from routinely collected electronic health record data[25,26] to minimize additional burden on doctors, nurses and patients.

Our results on the un-adjusted effect of the current FI (+3% per 0.01 FI) are highly similar to estimates from a recent study based on biannual longitudinal data from the US and Europe[16]. Our adjusted results, i.e. that the rate of change is a better predictor of mortality than the current FI, differ from what three previous studies found. An Australian[13] and a US[15] study found that both FI-change and the FI-level (after 4.5 years respectively 1 year of follow-up) were statistically significant predictors of mortality. In addition, Thompson et al.[13] also reported that the FI at follow-up was more predictive than at baseline. In a study based on three cohorts of older adults from Finland[17], Bai et al. recently found instead that the FI rate of change was no longer statistically significant after accounting for the current FI level. There are multiple potential explanations for these conflicting findings. First, the longer intervals between assessments (two or more years) in two of these studies[13,17] make it more difficult to capture FI changes adequately. Participants who experience substantial health deteriorations may drop out of the sample between assessments, thereby leading to an underestimation of FI increases, particularly in late-life[10]. Also, non-linear FI changes – e.g. an FI increase followed by a decrease due to a fall injury and subsequent recovery – cannot be captured adequately with only two FI measurement points[13,15]. Second, it could be due to age-related sample characteristics. We surveyed a sample of older adults that was several years older on average (82 years) – and hence more frail – than the participants in the other studies. Many older adults who survive into their 80s are multimorbid[28] and have accumulated a fair number of health deficits, yet their rate of accumulation is often relatively low[29]. Among the oldest old, recent frailty increases could reflect terminal health declines[10], and therefore be more predictive than (absolute) current frailty levels. This is also in line with the better predictive performance in 1-than 2-year mortality. Third, it also could be due to differences in methodology. Compared to Cox regression models, the joint models used in our analyses allow for a better monitoring of ups and downs in older adults frailty trajectories by allowing to take non-linear within-person changes into account, and by integrating participants with incomplete follow-up[23] data as well as considering measurement error[30] in FI estimates.

In conclusion, we found the rate of FI change to be more important than the current FI level for short-term mortality prediction among older adults 75+, which indicates a benefit of regular frailty assessments among this age group.

## Data Availability

Data from the Longitudinal Aging Study Amsterdam (LASA) are available for use for specific research questions, provided that an agreement is made up. Research proposals should be submitted to the LASA Steering Group, using a standard analysis proposal form that can
be obtained from the LASA website: www.lasa-vu.nl. Files with data published in this
publication are freely available for replication purposes and can be obtained using the same analysis proposal form. The LASA Steering Group will review all requests for data to ensure that proposals for the use of LASA data do not violate privacy regulations and are in keeping with informed consent that is provided by all LASA participants. The R-Markdown code reproducing all analyses, results and this manuscript are available
online (https://osf.io/8njg9/).

## Data and code availability

Data from the Longitudinal Aging Study Amsterdam (LASA) are available for use for specific research questions, provided that an agreement is made up. Research proposals should be submitted to the LASA Steering Group, using a standard analysis proposal form that can be obtained from the LASA website: www.lasa-vu.nl. Files with data published in this publication are freely available for replication purposes and can be obtained using the same analysis proposal form. The LASA Steering Group will review all requests for data to ensure that proposals for the use of LASA data do not violate privacy regulations and are in keeping with informed consent that is provided by all LASA participants.

The R-Markdown code reproducing all analyses, results and this manuscript are available online (https://osf.io/8njg9/).

## Author Contributions

E.S. planned the study, performed all statistical analysis, and wrote the article. H.M. contributed to the interpretation of results and critically reviewed the manuscript. E.O.H helped provide access to the data, helped with the construction of the frailty index, and critically reviewed the manuscript.

