## Appendix 1 for "Frailty trajectories in the oldest old: Is the level or the rate of change more predictive of mortality?"

**Supplementary data**

Health deficits included in the frailty index of the 75-PLUS-Study in LASA

|  | **Deficit** | **Cut-off values** |
| --- | --- | --- |
| 1 | Cardiac disease | No = 0, Yes = 1 |
| 2 | Peripheral arterial disease | No = 0, Yes = 1 |
| 3 | Stroke | No = 0, Yes = 1 |
| 4 | Diabetes | No = 0, Yes = 1 |
| 5 | Lung disease | No = 0, Yes = 1 |
| 6 | Osteoarthritis | No = 0, Yes = 1 |
| 7 | Rheumatoid arthritis | No = 0, Yes = 1 |
| 8 | Weight loss | No = 0, Yes = 1 |
| 9 | Pain while walking | No = 0, Yes = 1 |
| 10 | Constant pain | No = 0, Yes = 1 |
| 11 | Memory complaint | No = 0, Yes = 1 |
| 12 | Receiving help with domestic tasks | No = 0, Yes = 1 |
| 13 | Walk up/down staircase 15 steps without resting | Yes = 0, Yes, with some difficulty = 0.25, Yes, with much difficulty = 0.50, Only with help = 0.75, No = 1 |
| 14 | Dress/undress self | Yes = 0, Yes, with some difficulty = 0.25, Yes, with much difficulty = 0.50, Only with help = 0.75, No = 1 |
| 15 | Sit down/stand up from chair | Yes = 0, Yes, with some difficulty = 0.25, Yes, with much difficulty = 0.50, Only with help = 0.75, No = 1 |
| 16 | Cut own toenails | Yes = 0, Yes, with some difficulty = 0.25, Yes, with much difficulty = 0.50, Only with help = 0.75, No = 1 |
| 17 | Take shower | Yes = 0, Yes, with some difficulty = 0.25, Yes, with much difficulty = 0.50, Only with help = 0.75, No = 1 |
| 18 | Walk outside 5 minutes without stopping | Yes = 0, Yes, with some difficulty = 0.25, Yes, with much difficulty = 0.50, Only with help = 0.75, No = 1 |
| 19 | Use of transportation | Yes = 0, Yes, with some difficulty = 0.25, Yes, with much difficulty = 0.50, Only with help = 0.75, No = 1 |
| 20 | How is your health in general? | Excellent = 0, Good = 0.25, Fair = 0.50, Sometimes good/bad = 0.75, Poor = 1 |
| 21 | Feel depressed (CES-D) | Rarely or never = 0, Some of the time = 0.33, Occasionally = 0.66, Mostly or always = 1 |
| 22 | Feel everything is an effort (CES-D) | Rarely or never = 0, Some of the time = 0.33, Occasionally = 0.66, Mostly or always = 1 |
| 23 | Feel happy (CES-D) | Mostly or always = 0, Occasionally = 0.33, Some of the time = 0.66, Rarely or never = 1 |
| 24 | Feel lonely (CES-D) | Rarely or never = 0, Some of the time = 0.33, Occasionally = 0.66, Mostly or always = 1 |
| 25 | Restless sleep (CES-D) | Mostly or always = 0, Occasionally = 0.33, Some of the time = 0.66, Rarely or never = 1 |
| 26 | Could not get going (CES-D) | Rarely or never = 0, Some of the time = 0.33, Occasionally = 0.66, Mostly or always = 1 |
| 27 | Orientation time (MMSE) | Five correct = 0, One wrong = 0.50, Two or more wrong = 1 |
| 28 | Orientation place (MMSE) | Five correct = 0, One wrong = 0.50, Two or more wrong = 1 |
| 29 | Attention (MMSE) | Five correct = 0, One or two wrong = 0.50, Three or more wrong = 1 |
| 30 | Recall (MMSE) | Three correct = 0, Two correct = 0.50, One or zero correct = 1 |
| 31 | Gait speed (6 meters) | Normal = 0, Slow (>10 sec) or physically unable = 1 |
| 32 | Grip strength | Normal = 0, Weak = 1  Men:  BMI < 23.09 and GS <= 29 = 1  BMI >= 23.10 & BMI <= 26.09 and GS <= 30 = 1  BMI >= 26.1 & BMI <= 28 and GS <= 30 = 1  BMI >= 28.01 and GS <= 32 = 1  Women:  BMI < 23.09 and GS <= 17 = 1  BMI >= 23.10 & BMI <= 26.09 and 17.3 <= 30 = 1  BMI >= 26.1 & BMI <= 28 and GS <= 18 = 1  BMI >= 28.01 and GS <= 21 = 1 |

All health deficits had <5% missing values, and the FI was calculated only for participants who provided valid information for at least 80% of the health deficits, which applied to >99% in each wave.
